# Relationship Between Obesity and Balance in Parachinar, District Kurram, Kpk-An Analytical Cross-Sectional Study

**DOI:** 10.1101/2023.03.18.23287439

**Authors:** Sayed Sajid Hussain

**Affiliations:** Sarhad University of Sciences and information Technology, Peshawar; Combined Military Hospital (CMH) Peshawar

**Keywords:** Obesity, Falls, BBS, BMI TUG

## Abstract

**OBJECTIVE:** To find out the effects of BMI on BBS and to determines the risk of falls due to impaired balance in Parachinar, District Kurram

**METHODS:** An analytical cross-section study was carried out to determine the effects of BMI on BBS; the study included 269 individuals who are classified based on their body mass index. There were 157 males and 112 female. The clinical evaluation instruments Berg balance scale and Time Up and Go test (TUG) were used to collect the data and investigate the relationship between body mass index-based obesity and balance. Data was collected in Parachinar District Kurram from individuals have BMI 22.5kg/m2 or greater and age 20-50 years.

**RESULTS:** The BBS and TUG scores revealed that obese people had poor balance due to their body weight, as well as a decline in body balance and stability performance. There were 71.4% individuals who had BMI of 27.5-32.5kg/m^2^ (class 1 obesity) and 10% individuals had severed obesity (class 3). 66.9% individuals had low risk of fall, 20% had medium risk of fall, and 12.3% had high risk of fall.

**CONCLUSION:** This study concluded that obesity has effects on balance and obese individuals have poor balance performance. According to this study individuals with greater BMI have poor balance performance and have high risk of falls. The results of this study illustrates that there is an association of BMI and BBS because the P-value is <0.05 and concluded that obesity cause impaired balance and which leads to increase a risk of falls.

## INTRODUCTION

Obesity can also be defined as a preventable medical condition associated with the accumulation of excessive fat due to genetic and environmental factors [1], or obesity can also be defined as abnormal and excessive fat accumulation with a body mass index (BMI) of 25kg/m2 or higher for Asian people [2], who have a higher proportion of fat and a higher BMI (BMI)..Obesity and overweight put a constant strain on the planter mechanoreceptors, resulting in decreased planter sensitivity and poor and compromised balance [2,3]. When we compare the abilities of obese people to those of healthy people, we find that the functional abilities of the obese people are lower [3]. There is a significant difference between obesity and overweight. The World Health Organization (WHO) distinguishes obesity and overweight based on body mass index (BMI) [3,4]. Overweight signifies an excess of body weight, whereas obesity denotes an excess of body fat [4]. In Asia, a BMI of 24.9 - 27kg/m2 is considered overweight, while a BMI of 27.5kg/m2 or above is considered obese, according to the International Association for the Study of Obesity Task Force [5].

Obesity causes a variety of medical disorders, including heart disease, diabetes, cancer, respiratory problems, musculoskeletal ailments, and poor balance [6]. Obesity is linked to a loss of postural control and stability, as well as a reduction in bone density and joint function. Obesity has an impact on walking speed and patterns, and obese persons walk slower than healthy people due to their slower velocity and shorter stride length [7]. Balance is defined as the capacity to keep one’s centre of mass within a stable base of support, which is necessary for normal upright movement and physical activity [8]. Balance refers to an individual’s capacity to maintain proper vision during dynamic posture, recognize the speed and direction of movement, maintain posture, and offer stability in a variety of situations [8,9]. Maintaining balance is a complicated process involving multiple components, including the sensory and motor systems, which are in charge of body feeling and movement, and the higher level of the CNS, which is in charge of processing sensory data and issuing commands or initiating movement [10]. Sensory system, proprioceptive and vestibular system is the most common factors that contribute to achieve and maintain balance [10,11]. Both obesity and body stability offers the effort to bring the static and dynamic body close to equilibrium in vary conditions [12]. Several studies show that body mass index (BMI) has directly relationship with balance in static position as well as dynamic position but the category is not yet clear for physical activities and balance [13]. Physical activities (PA) at a higher level necessitate a higher degree of balance, hence physical activities and body mass index (BMI) have an impact on balance [12,13]. In a study which was conducted by Pollack and Chestin showed that obese people have higher risks of knee, hand, fingers, and wrist injuries when compared to healthy people [14]. Obesity can alter balance especially in dynamic postural stability, and has effects on calf muscle stretching and alters kinematics of ankle joint, and obese individuals are at greater risk for developing mobility limitations and older obese individuals the limitations and impairments of mobility seen greater and many falls are the result of impaired balance [15,16]. When compares the ability of prolonged physical activities in obese and non-obese there will be deterioration of postural control and at high risk for fall [15], according to Fretitas *et al* (2005) during 30 minutes of prolonged standing tasks, it is noted incensement in postural sway in elderly and younger. Studies show that balance is altered when BMI increases and it is noted especially in dynamic posture at different dynamic stability level. According to Petrolimi *et al* obesity impacts the sensory motor and central nervous system (CNS) which are responsible for maintain balance during dynamic and static postures in different circumstances. Obesity and overweight progressively put pressure on plantar mechanoreceptors due to which obese individuals have poor and impaired balance therefore if we compare the functional abilities of normal individuals with obese there will be decline in balance performance [16]. Obesity is the risk factor for reducing postural control and stability, and obesity has negative effects on bone density and joint functions. Obesity has also linked with the walking speed and patterns of individual’s therefore obese individuals generally have slow walking speed if we compare healthy individuals because obese individuals have slower velocity and shorter stride length [17]. The impaired balance due to obesity is the main risk of fall because obesity declines the performance of body balance which leads to causes falls [17]. Falls are defined as the accidental events where one’s lost his/her center of gravity and decline occurs in balance, where no efforts are made to restore or maintain balance or the applied efforts are not much enough to maintain balance [18]. The incidence and severity of fall and its related complications are directly proportion to increasing age, obesity, and impaired balance therefore obese older individuals are more prone to falls than younger ones due to physiological declination related to age and declines in balance and there are numerous factors that are contributing to falls but two are important to assess and have been identified: muscle weakness and impaired balance [19]. According to data, falls are the second leading cause of mortality worldwide. The 2010 Global Burden of Disease and Injury study showed that 12% of all unintentional injury death is due to falls [18,19]. Falls can lead to significant disability especially in older population that requires prolong treatment [20].

## METHODS

The study design was an analytical cross-sectional study (November 2022 to April 2022). After approval of Advance Studies and Research Board (ASRB), graduate committee and Ethical board of Sarhad University of Science and Information Technology the data was collected. The sample size calculated for this study was 269 through OpenEpi. Non-probability Convenience sampling technique was used and data was analyzed by using software SPSS 25.

### Inclusion Criteria

- Individuals without recent trauma and don’t have any fracture.
- Individuals that is independent and mobile without assisted devices.
- Both male and female of different ages ranges from 20-50 year
- Individuals have BMI 27.5kgm2 or above

### Exclusion Criteria

- Individuals have visual, vestibular insufficiency, and recent trauma.
- Individuals who have not independently mobility.
- Known acute illness and current fracture.
- Medication that affects muscle strength and balance (Steroids)
- Cancer (Malignancy, Chemotherapy)
- Musculoskeletal and Neurological diseases like Parkinson’s and stroke.

### DATA COLLECTION PROCEDURE AND PERMISSION TAKEN FROM SUIT REASEARCH AND ETHICAL COMITTE

Standard medical tools for examination of balance (Berg balance scale and TUG) were used to calculate the obese individuals balance and find out the risk of falls. The data collected from Parachinar District Kurram, KPK. Before data collection, I took approval from the head of the orthopedic department of District Head Quarter Hospital Parachinar, KPK. Then take written consent from the individuals who show willingness to participate in the current study and then they were screened for inclusion and exclusion criteria. An adopted questionnaire from the literature was used in the study which was completed by the Physical therapists working in neurological departments of their respective hospitals. All the information regarding questionnaire were shared with the participants who were include questions about the demographic of the participants, Berg balance and TUG scores respectively.

## RESULTS

In this study there were 269 participants, there were 157 were males and 112 were females. Out of 269, there were 156 (58%) individuals with aged 20-25 year, 86(32%) with aged 26-30 year, 17(6.3%) with aged 31-35 year, and 10 (3.7%) with aged 36-40 year. Among 269 participants there were 192(71.4%) participants with BMI 27.5-32.5kg/m^2^, 50(18.6%) participants with BMI 32.5-37.5kg/m^2^, and only 27(10%) with BMI 37.5/m^2^ or above. The sever type of obesity (Class 3 obesity) noted only in 10% respondents. Out of 269 participants, 150 (55.8%) participants scored in between 41-56, 92 (34.2%) participants scored in between 21-40, and 27 (10%) participants scored in between 0-20. These values are used for the predication of falls lower the BBS greater the chance of falls and high impaired balance. According to the figure 1.4 55.8% participants indicates low impaired balance while 10% shows greater impaired balance on the basis on BBS. 242 participants out of total 269 have TUG score ≤20 seconds which indicates good mobility, can go alone, without use of gait aid and 27 out of total 269 have TUG score ≤30 indicates problems, cannot go outside alone, requires care and gait aid.

Table 2 illustrates that there is association of BMI and BBS when apply Chi-square test as the P-value is <0.05 which shows that obese individuals have impaired balance. Furthermore as the BBS decreases the mean value is increased which shows the relationship of fall due to impaired balance recorded by BBS. The mean value for low fall risk is 1.000 while, It increases as BBS increase.

| Age | Frequency | Percentage |
| --- | --- | --- |
| 20-25 year | 156 | 58.0 |
| 26-30 year | 86 | 32.0 |
| 31-35 year | 17 | 6.3 |
| 36-40 year | 10 | 10.0 |

| BMI | Frequency | Percentage |
| --- | --- | --- |
| **Class 1: BMI of 27.5 to 32.5kg/m<sup>2</sup>** | 192 | 71.4 |
| **Class 2: BMI of 32.5-37.5kg/m<sup>2</sup>** | 50 | 18.6 |
| **Class 3 : BMI of 37.5kg/m<sup>2</sup>** | 27 | 10.0 |
| Respondent BBS | Frequency | Percentage |
| **Obtained Score 41-56** | 150 | 55.8 |
| **Obtained Score 21-40** | 92 | 34.2 |
| **Obtained Score 0-20** | 27 | 10.0 |
| Risk of fall | Frequency | Percentage |
| **low fall risk** | 180 | 66.9 |
| **medium fall risk** | 56 | 20.8 |
| **high fall risk** | 33 | 12.3 |

**Table 2:** shows P-value and association

| Chi-Square Tests | Value | Df | Asymp. Sig. |
| --- | --- | --- | --- |
| **Pearson Chi-Square** | 445.886<sup>a</sup> | 4 | .003 |
| **Likelihood Ratio** | 352.470 | 4 | .000 |
| **Linear-by-Linear Association** | 210.662 | 1 | .004 |

## DISCUSSION

The aim of the current research study was to find out the weather body mass-based obesity has effects on balance performance and identifies the risk of falls based on BBS. For this purpose, I took data from adults of age between 20 to 40 years in which there were 157(58.4%) male and 122(41.6%) female. Among these there were obesity class I in 71.4%, obesity class II in 18.6% and obesity class III in 10% of respondents. There was low fall risk in 180(66.9%), medium fall risk in 56(20.8%), and high fall risk in 33(12.3%) individuals. The results of this study illustrates that there is an association of BMI and BBS because the P-value is <0.05 and concluded that impaired balance due to obesity is a risk of falls. A study by Jose M et al. in 2018 found that there is association of BMI with static and dynamic balance because the P-value was <0.05 and concluded that obesity is a risk factor for fall due to impaired balance, so the current study has also P-value <0.05 shows that and because in obese individuals there is declines in balance due to over pressurizing of planter mechanoreceptors and body weight, load, and stress also affects balance performance [16]. A study in 2015 done by Marcose Rossi et al. found that obese individuals took longer duration to perfume modified TUG and need more steps to complete and the P-value was <0.048, so the current research study reveals participants with greater BMI took longer time to complete TUG and P-value is <0.05 but the age of participants are different. The P-value determined by them was <0.04 while in the current research study the P-value is 0.000. Obesity affects all aged people in Pakistan especially adults in urban area, so the current study focuses in adults and concludes that there is association of BMI and TUG performance [15]. Obese group individuals have slower velocity and short stride length due to which they took longer time to perfume TUG [17]. A study in 2015 done by Marcose Rossi et al. found that obese individuals took longer duration to perfume modified TUG and need more steps to complete and the P-value was <0.048, so the current study shows that participants with greater BMI took longer time to complete TUG and P-value is <0.05 but the age of participants are different. The P-value determined by them was <0.04 while in the current research study the P-value is 0.000. Obesity affects all aged people in Pakistan especially adults in urban area, so the current study focuses in adults and concludes that there is association of BMI and TUG performance. Obese group individuals have slower velocity and short stride length due to which they took longer time to perfume TUG [19].

## CONCLUSION

This study concluded that there is relationship between obesity and balance on the basis of statistical analysis. This study has revealed that individuals with greater BMI have poor balance performance and have high risk of falls. Most of participants have obesity class I and aged individuals have low score of BBS and high risk of falls due loss of balance parameter and stability.

## Data Availability

All data produced in the present study are available upon reasonable request to the authors

