## Appendices for "Relationship Between Obesity and Balance in Parachinar, District Kurram, Kpk-An Analytical Cross-Sectional Study"

**Appendix 1**

**RELATIONSHIP BETWEEN OBESITY AND BALANCE IN PARACHINAR, DISTRCT KURRAM**

**Name: __________________________________**	**Dat**e: ___________________

**Location: ________________________________**	**Rater: ___________________**

**Weight: _________________________________	Height: ___________________**

**Body Mass Index (BMI**

**BMI = ________kg/m2**

**Obese class: _________**

- The healthy range is 18.0 - 22.9kg/m^2^ .BMI applies to most adults 18-65 years.
- A BMI of 23.0 – 27kg/m^2^  or more is overweight
- Obese class I = 27.5 - 32.5kg/m^2^
- Obese class II = 32.5-37.5kg/m^2^
- Obese class III= 37.5kg/m^2^  or above

**Berg Balance Scale**

**ITEM DESCRIPTION	SCORE (0-4)**

1. Sitting to standing	________
2. Standing unsupported	________
3. Sitting unsupported	________
4. Standing to sitting	________
5. Transfers	________
6. Standing with eyes closed	________
7. Standing with feet together	________
8. Reaching forward with outstretched arm	________
9. Retrieving object from floor	________
10. Turning to look behind	________
11. Turning 360 degrees	________
12. Placing alternate foot on stool	________
13. Standing with one foot in front	________
14. Standing on one foot	________

**Total Score ________**

- A score of 56 indicates functional balance
- A score of < 45 indicates individuals may be at greater risk of falling
- 41-56 = low fall risk
- 21-40 = medium fall risk
- 0-20 = high fall risk

**Category_____________**

**Berg Balance Scale**

**1. SITTING TO STANDING**

INSTRUCTIONS: Please stand up. Try not to use your hand for support.

(4) Able to stand without using hands and stabilize independently

(3) Able to stand independently using hands

(2) Able to stand using hands after several tries

(1) Needs minimal aid to stand or stabilize

(0) Needs moderate or maximal assist to stand

**2. STANDING UNSUPPORTED**

INSTRUCTIONS: Please stand for two minutes without holding on.

(4) Able to stand safely for 2 minutes

(3) Able to stand 2 minutes with supervision

(2) Able to stand 30 seconds unsupported

(1) needs several tries to stand 30 seconds unsupported

(0) unable to stand 30 seconds unsupported

If a subject is able to stand 2 minutes unsupported, score full points for sitting unsupported. Proceed to item #4.

**3. SITTING WITH BACK UNSUPPORTED BUT FEET SUPPORTED ON FLOOR OR ON A STOOL**

INSTRUCTIONS: Please sit with arms folded for 2 minutes.

(4) Able to sit safely and securely for 2 minutes

(3) Able to sit 2 minutes under supervision

(2) Able to able to sit 30 seconds

(1) Able to sit 10 seconds

(0) unable to sit without support 10 seconds

**4. STANDING TO SITTING**

INSTRUCTIONS: Please sit down.

(4) Sits safely with minimal use of hands

(3) Controls descent by using hands

(2) Uses back of legs against chair to control descent

(1) Sits independently but has uncontrolled descent

(0) needs assist to sit

**5. TRANSFERS**

INSTRUCTIONS: Arrange chair(s) for pivot transfer. Ask subject to transfer one way toward a seat with armrests and one way toward a seat without armrests. You may use two chairs (one with and one without armrests) or a bed and a chair.

(4) Able to transfer safely with minor use of hands

(3) Able to transfer safely definite need of hands

(2) Able to transfer with verbal cuing and/or supervision

(1) Needs one person to assist

(0) needs two people to assist or supervise to be safe

**6. STANDING UNSUPPORTED WITH EYES CLOSED**

INSTRUCTIONS: Please close your eyes and stand still for 10 seconds.

(4) Able to stand 10 seconds safely

(3) Able to stand 10 seconds with supervision

(2) Able to stand 3 seconds

(1) Unable to keep eyes closed 3 seconds but stays safely

(0) Needs help to keep from falling

**7. STANDING UNSUPPORTED WITH FEET TOGETHER**

INSTRUCTIONS: Place your feet together and stand without holding on.

(4) Able to place feet together independently and stand 1 minute safely

(3) Able to place feet together independently and stand 1 minute with supervision

(2) Able to place feet together independently but unable to hold for 30 seconds

(1) Needs help to attain position but able to stand 15 seconds feet together

(0) Needs help to attain position and unable to hold for 15 seconds

**8. REACHING FORWARD WITH OUTSTRETCHED ARM WHILE STANDING**

INSTRUCTIONS: Lift arm to 90 degrees. Stretch out your fingers and reach forward as far as you can. (Examiner places a ruler at the end of fingertips when arm is at 90 degrees. Fingers should not touch the ruler while reaching forward. The recorded measure is the distance forward that the fingers reach while the subject is in the most forward lean position. When possible, ask subject to use both arms when reaching to avoid rotation of the trunk.)

(4) Can reach forward confidently 25 cm (10 inches)

(3) Can reach forward 12 cm (5 inches)

(2) Can reach forward 5 cm (2 inches)

(1) Reaches forward but needs supervision

(0) loses balance while trying/requires external support

**9. PICK UP OBJECT FROM THE FLOOR FROM A STANDING POSITION**

INSTRUCTIONS: Pick up the shoe/slipper, which is place in front of your feet.

(4) Able to pick up slipper safely and easily

(3) Able to pick up slipper but needs supervision

(2) Unable to pick up but reaches 2-5 cm(1-2 inches) from slipper and keeps balance independently

(1) Unable to pick up and needs supervision while trying

(0) Unable to try/needs assist to keep from losing balance or falling

**10. TURNING TO LOOK BEHIND OVER LEFT AND RIGHT SHOULDERS WHILE STANDING**

INSTRUCTIONS: Turn to look directly behind you over toward the left shoulder. Repeat to the right. Examiner may pick an object to look at directly behind the subject to encourage a better twist turn.

(4) Looks behind from both sides and weight shifts well

(3) Looks behind one side only other side shows less weight shift

(2) Turn sideways only but maintain balance

(1) Needs supervision when turning

(0) Needs assist to keep from losing balance or falling

**11. TURN 360 DEGREES**

INSTRUCTIONS: Turn completely around in a full circle. Pause. Then turn a full circle in the other direction.

(4) Able to turn 360 degrees safely in 4 seconds or less

(3) Able to turn 360 degrees safely one side only 4 seconds or less

(2) Able to turn 360 degrees safely but slowly

(1) Needs close supervision or verbal cuing

(0) needs assistance while turning

**12. PLACE ALTERNATE FOOT ON STEP OR STOOL WHILE STANDING UNSUPPORTED**

INSTRUCTIONS: Place each foot alternately on the step/stool. Continue until each foot has touch the step/stool four times.

(4) Able to stand independently and safely and complete 8 steps in 20 seconds

(3) Able to stand independently and complete 8 steps in > 20 seconds

(2) Able to complete 4 steps without aid with supervision

(1) Able to complete > 2 steps needs minimal assist

(0) needs assistance to keep from falling unable to try

**13. STANDING UNSUPPORTED ONE FOOT IN FRONT**

INSTRUCTIONS: (DEMONSTRATE TO SUBJECT) Place one foot directly in front of the other. If you feel that you cannot place your foot directly in front, try to step far enough ahead that the heel of your forward foot is ahead of the toes of the other foot. (To score 3 points, the length of the step should exceed the length of the other foot and the width of the stance should approximate the subject’s normal stride width.)

(4) Able to place foot tandem independently and hold 30 seconds

(3) Able to place foot ahead independently and hold 30 seconds

(2) Able to take small step independently and hold 30 seconds

(1) Needs help to step but can hold 15 seconds

(0) loses balance while stepping or standing

**14. STANDING ON ONE LEG**

INSTRUCTIONS: Stand on one leg as long as you can without holding on.

(4) Able to lift leg independently and hold > 10 seconds

(3) Able to lift leg independently and hold 5-10 seconds

(2) Able to lift leg independently and hold ≥ 3 seconds

(1) Tries to lift leg unable to hold 3 seconds but remains standing independently.

(0) unable to try of needs assist to prevent fall

**Total score** ________

**Timed Up and Go Test (TUG)**

**Instruction**

1. Patients wear their regular footwear and can use a walking aid, if needed.
2. The patient starts in a seated position
3. The patient stands up upon therapist’s command: walks 3 meters, turns around, walks back to the chair and sits down.
4. The time stops when the patient is seated.
5. Be sure to document the assistive device used.

- ≤10 seconds = normal
- ≤20 seconds = good mobility, can gout alone, mobile without gait aid
- ≤30 seconds = problems, cannot go outside alone, requires gait aid
- A score of ≥14 seconds has been shown to indicate high risk of falls

**Total score** __________

**Consent form**

**Appendix 2**

**RELATIONSHIP BETWEEN OBESITY AND BALANCE IN PARACHINAR, DISTRCT KURRAM**

Participant name _______________

Age_________

Gander_________

Cell No___________________

Address_______________________________________________

Title of Project: RELATIONSHIP BETWEEN OBESITY AND BALANCE IN PARACHINAR, DISTRCT KURRAM

Name of researcher

Sayed Sajid Hussain

Please pick all the boxes

1. I confirmed that I have read and understand the objective and aim of the study for the above study.
2. I understand that my participation in the study is voluntarily.
3. I agree to take part in the above research study.

Signature of Participant ___________________
